# Improving PEG Tube Management in ALS Patients Through Educational Pamphlets

**DOI:** 10.64898/2025.12.09.25336480

**Authors:** Minh Nguyen, Matthew Penna, Mitchell Powell, Kevin Pallapati, Lydia Sharp, James Peter Orengo

**Author notes:** **Funding:** None.

## Abstract

Patients with amyotrophic lateral sclerosis (ALS) often experience dysphagia, leading to malnutrition and weight loss. Percutaneous endoscopic gastrostomy (PEG) tubes help address these issues, but complications and limited patient education may diminish their benefits. This project evaluated the patient experience from an educational pamphlet created by a multi-disciplinary team at our academic center’s ALS Clinic. The pamphlet was distributed at the time of PEG placement and covered tube function, maintenance, and troubleshooting. A retrospective survey compared the experiences of patients who received the pamphlet to a control group of patients who did not. Among 22 respondents, 45% received the pamphlet. Pamphlet users reported better preparedness (100% vs. 58%), confidence in PEG care (90% vs. 58%), and pre-placement education (90% vs. 42%). Overall satisfaction with the PEG tube was high (82%), although satisfaction with the placement process was lower (68%). These findings informed us that accessible educational interventions may improve patient outcomes and experiences in our ALS patient population.

## Main Text Introduction

Patients with amyotrophic lateral sclerosis (ALS) commonly experience dysphagia, leading to malnutrition and weight loss. Percutaneous endoscopic gastrostomy (PEG) tubes are used to maintain long-term nutrition in ALS patients to address these challenges. However, we noted in our multi-disciplinary ALS clinic that often patients and caregivers face challenges with PEG tube maintenance due to limited education, particularly in dealing with complications. Common complications of PEG tubes include dislodgement, site infection, and clogging, all of which may diminish the benefit of PEG tubes and increase caregiver burden.^1^ While multi-disciplinary ALS clinics aim to provide comprehensive counseling, patients and caregivers frequently report gaps in procedural education and support.

Structured education using verbal, written, and visual tools has been shown to improve outcomes in home enteral nutrition and is recommended to begin prior to PEG placement.^2,3^ In both ALS and broader neuromuscular populations, the use of standardized pamphlets, checklists, and demonstrations has been associated with increased caregiver competence, reduced complications, and improved adherence to care routines.^4^

To address unmet educational needs in PEG tube management, the goal of this quality improvement project was to develop and implement an educational pamphlet for patients undergoing PEG placement and assess its impact on patient-reported measures of preparedness, confidence, and satisfaction.

## Methods

A multi-disciplinary team at our academic center’s ALS clinic designed a user-friendly pamphlet addressing PEG tube function, maintenance, troubleshooting, instructions on when to seek medical attention, and frequently asked questions. Beginning in January 2021, the pamphlet was distributed to patients and caregivers prior to PEG tube placement. A retrospective survey including Likert-scale and open-ended questions was administered to patients who received the pamphlet and a comparison group that did not. Baseline demographics of survey respondents are shown in Table 1. Survey questions assessed preparedness, confidence, and satisfaction with PEG tube use and education. Patients and caregivers were also asked to identify specific challenges with PEG tube maintenance.

**Table 1.**
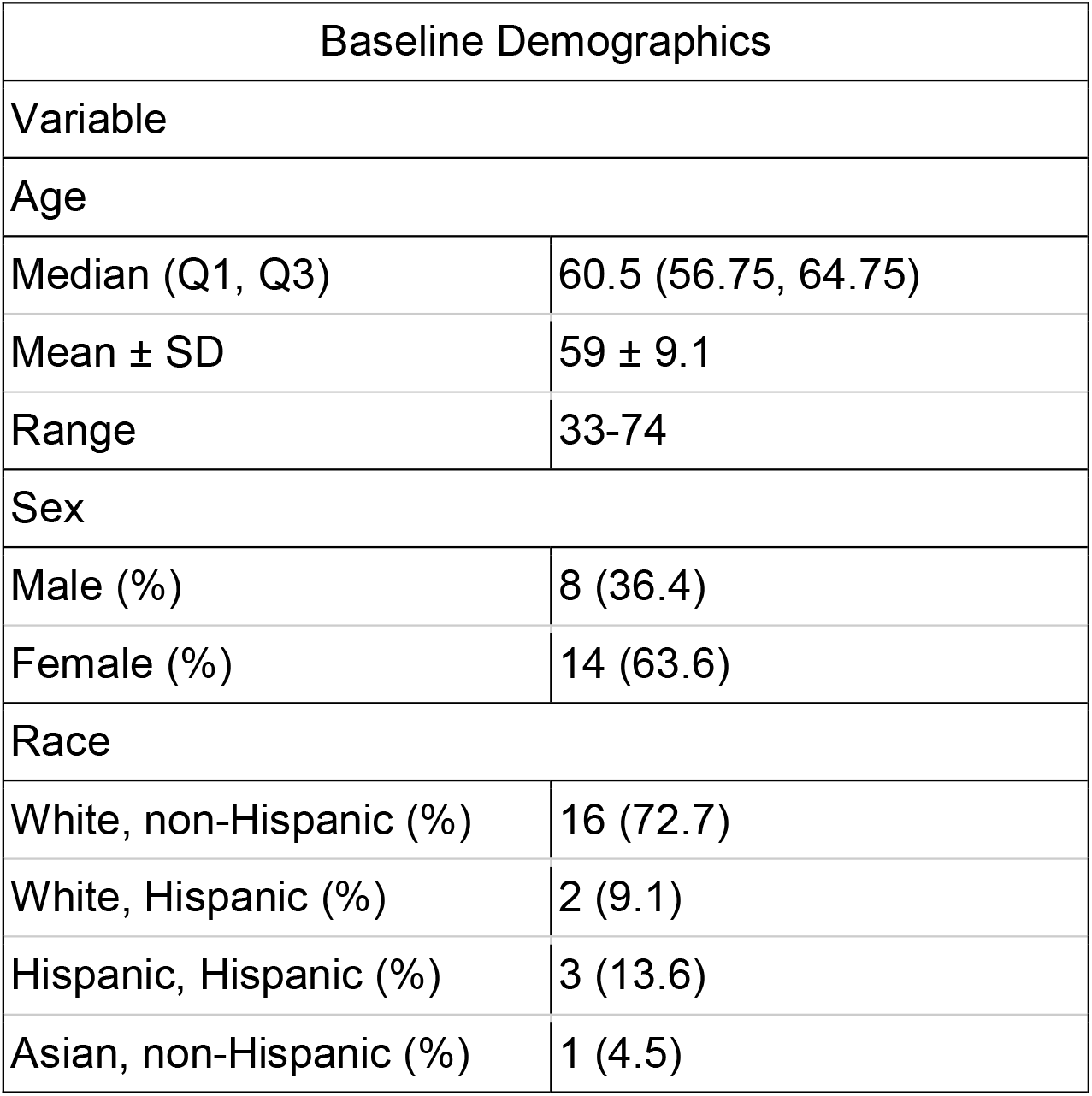
Baseline Demographics of Survey Respondents.

## Results

Among 22 survey respondents, 45% (10/22) reported receiving and using the pamphlet. Compared to those who did not use the pamphlet, pamphlet users reported higher preparedness in management (100% vs. 58%), confidence in PEG care (90% vs. 58%), and better pre-placement education (90% vs. 42%) compared to non-users.

Overall, 82% of respondents were satisfied with the PEG tube, though satisfaction with the placement process was lower (68%). Half of the respondents identified specific challenges with PEG tube use or the placement process, including complications with PEG tube use, limited access to maintenance equipment, and difficulties contacting the care team for follow-up when issues arose.

Pamphlet users were more likely to report a positive experience and cited the resource as helpful for reinforcing clinic-based education. Non-users more often described feeling unprepared or receiving inadequate education.

## Discussion

These findings suggest that low-cost, educational interventions may improve patient outcomes in the ALS population. Patients who received and utilized the pamphlet at or site felt more prepared, confident, and educated regarding the maintenance and use of their PEG tubes, highlighting the importance of supplemental materials in supporting clinic counseling.

These results support current recommendations from clinical nutrition societies, which advocate for the initiation of structured education prior to the placement of PEG tubes to promote home care readiness.^2,3^ Similar interventions, including printed guides, video demonstrations, and live caregiver training, have been shown to reduce PEG tube complications and improve caregiver confidence.^4^ These strategies are especially beneficial in the ALS population, where patients and caregivers often face enhanced care demands and emotional strain.

Limitations of the project include a small sample size and potential recall bias. Self-reported measures do not capture objective complication rates. Future directions can focus on assessing how educational resources impact clinical outcomes, such as tube-related infections and nutritional metrics. Additionally, supplementing printed resources with digital tools or in-person demonstrations may further enhance retention and patient experience.

## Data Availability

All non PHI data produced in the present study are available upon reasonable request to the authors

## Acknowledgments

We thank the patients and caregivers of the Baylor College of Medicine ALS Clinic for their participation and feedback.

## Data Availability Statement (DAS)

Data not included in the article due to space limitations can be shared in anonymized form upon request by any qualified investigator for purposes of replicating procedures and results.

